# Early Onset Mental Health Problems, Educational Attainment and Productivity Loss in England: Evidence from the Millennium Cohort Study

**DOI:** 10.64898/2026.05.31.26354541

**Authors:** Shuye Yu, Jack Pollard, Tessa Reardon, Cathy Creswell, Ruth Wadman, Mara Violato

## Abstract

Mental health problems, including emotional problems, are linked to adverse educational outcomes among children and adolescents. This study examines the association between early onset of mental health problems generally, and emotional problems specifically, at ages 5-14, and outcomes from the General Certificate of Secondary Education (GCSE), a secondary education qualification, at age 16 for 4,783 students in England, using the Millennium Cohort Study dataset linked to the National Pupil Database. We found that the onset of mental health problems at ages 5, 7, 11 and 14 had a significant and negative association with all GCSE outcomes. We also found negative associations between early onset emotional problems and GCSE outcomes, although results were most stark for emotional problems that onset at age 11, with statistically significant negative associations with all GCSE outcomes. School absence was identified as a potential mediator of the negative association. Furthermore, this study found that the potential loss of productivity related to mental health problems in general and emotional problems in particular was over £23,000 and £11,000 per affected individual, respectively, which could translate into approximately £2.57 billion and £1.6 billion, respectively at the population level for England. These findings highlight the importance of early intervention for children and adolescents with mental health problems to improve educational and future outcomes.

## 1. Introduction

The rising global prevalence of mental health problems in children and young people is a critical public health concern (Benton et al., 2021). In England, a national survey conducted in 2023 reported that 20.3% of children and young people aged 8-16 years were identified as having a probable mental health disorder, revealing a sharp increase from 12.5% in 2017 (NHS England, 2023). Among the various mental health problems affecting this age group, emotional problems, i.e. anxiety disorders and depression, are often the most common (Kieling et al., 2024; Polanczyk et al., 2015). Existing evidence indicates that emotional problems also have a particularly early age of onset, with a peak age of onset for anxiety disorders as early as 5.5 years (Solmi et al., 2022).

The early onset of mental health problems in general, and emotional problems in particular, can have persistent and multifaceted impacts across the life course. For example, a growing, but still embryonic, body of evidence underlines the long-term negative outcomes specifically associated with childhood anxiety problems, including persistence of mental health problems into adulthood, poor physical health, reduced educational attainment, lower employment, diminished financial well-being (Pollard et al., 2023), as well as worse health-related quality of life and increased healthcare costs (Dona et al., 2025; Pollard et al., 2023). As disruption to education often emerges at a younger age than other negative outcomes (e.g. reduced employment and financial wellbeing), educational outcomes warrant particular attention, especially given that academic attainment, and specifically success in some high-stakes examinations, plays a crucial role in both higher education (Jerrim, 2023) and future labour market engagement (Anderson, 2023).

In the education system of England, the General Certificate of Secondary Education (GCSE) attainment (details available in Appendix A1), is a critical milestone and is widely used as an indicator of educational performance at age 16 (Crawford, 2014). Achieving five or more GCSE subjects at grades A*– C (or 9 – 4 under the new grading system), including English and Mathematics, represents the basic threshold for further secondary education and most university programmes. More selective universities usually require significantly higher grades. Consequently, poor GCSE performance, can diminish access to higher education, ultimately resulting in lost opportunities for human capital development (Card, 1999).

Furthermore, the relationship between educational attainment and earnings (Card, 1999, 2001; Heckman et al., 2018) is well documented, including in the context of the GCSE qualification specifically (Hodge et al., 2021; Mcintosh, 2006). This means that the association between mental health problems and GCSE performance can be translated into measurable productivity losses.

Despite the early onset of mental health problems generally, and emotional problems specifically, studies have tended to focus on educational attainments of young people who experienced mental health problems aged 14 – 18 years (Butterworth & Leach, 2018; Cornaglia et al., 2015; Moore et al., 2020; Zając et al., 2024), with only a few focusing on cohorts who experienced mental health problems before age 14 (Currie et al., 2014; Dalsgaard et al., 2020; van Poortvliet, 2024). Further longitudinal evidence to explore the educational outcomes associated with early onset mental health problems is clearly required, with consideration of likely mechanisms driving any associations. A key candidate mechanism through which early mental health problems may affect educational outcomes is school absenteeism. Notably, anxiety disorders, across ages 5 to 21 years, have been shown to be positively associated with multiple forms of school absenteeism, including excused/medical absences, truancy, and school refusal (Finning et al., 2019). Such patterns of absence are likely to disrupt learning continuity, weaken school engagement, and lead to academic underperformance.

There is also a still limited body of literature exploring the relationship between mental health problems and aspirations or motivation to attend university. For example, a large population-based study of Norwegian adolescents found that higher levels of social anxiety at ages 13–15 years, measured using both self-reported instruments and diagnostic interviews, were associated with lower odds of aspiring to higher education (Jystad et al., 2021).

This study examined the association between early onset mental health problems (age 5–14 years) and GCSE performance, and explored whether any significant associations were mediated by school absenteeism. We used two indicators of mental health problems: (i) presence of mental health problems in general (using a broad measure of emotional and behavioural symptoms) and (ii) presence of emotional problems (using a measure of emotional symptoms) specifically, and we separately examined the first onset at ages 5, 7, 11 and 14. The analyses leveraged a linked dataset composed of the Millennium Cohort Study, which has tracked children born in the UK at the start of the 21st century (2000/02) up to age 23, and the National Pupil Database, which provides a rich administrative record of students’ school performance, including GCSE results. We also explored the potential labour income and productivity loss, at the individual and societal levels, associated with early-life mental health and emotional problems. Finally, although our data do not include a direct measure of educational aspirations, we use both child- and parent-reported perceived likelihood of attending university to explore the relationship between mental health problems and expectations regarding higher education.

## 2. Methods

### 2.1 Data and Sample Construction

We used data from the Millennium Cohort Study (MCS) (Hansen, 2014), a nationally representative birth cohort study of 18,818 children, from 18,552 families, born in the UK from 2000 to 2002. The MCS collects a variety of information on children and their parents/carers, including their health, sociodemographic characteristics, and a variety of family circumstances. Children have to date been followed through eight surveys, at ages 9 months, and 3, 5, 7, 11, 14, and 17 years, with age 23 recently released (but not included in this study). MCS data can be linked to the National Pupil Database (NPD) (University College London et al., 2022), an administrative education record held by the Department for Education in England, which comprises an extensive set of educational outcomes for pupils in state-funded schools in England, including GCSE results. Since GCSE exams take place at age 16, we only included children for whom mental health status was available and consistently reported by one of their biological parents, at ages 5, 7, 11, and 14 years. The sample was further restricted to those children who could successfully be matched to the NPD to obtain their GCSE outcomes at age 17 years. This meant our sample was restricted to children registered at schools in England, and excluded those in Scotland, Wales, Northern Ireland. Inclusion/exclusion criteria are reported in Appendix Figure A1.

### 2.2 Main Exposure: Mental Health Problems

The main exposure of interest was the mental health status of children, and we were particularly interested in the association between onset of mental health problems and educational outcomes for different ages of onset. We referred to the first appearance of mental health problems as a mental health “shock”. We focused on two aspects of mental health: (i) presence of mental health problems in general (henceforth called overall mental health problems – Exposure 1), and (ii) presence of emotional problems specifically (Exposure 2). Both exposures were derived from the parent-reported *Strengths and Difficulties Questionnaire* (SDQ), a brief emotional and behavioural screening tool that is widely used in general population studies to identify mental health problems in children and young people aged 2-17 years (Goodman, 1997, 1999, 2001). The SDQ comprises 25 questions, each measuring a different psychological attribute on a 3-point Likert scale. The SDQ items are grouped into five sub-scales, each containing five questions to assess: emotional symptoms; conduct problems; hyperactivity/inattention; peer relationship problems; and pro-social behaviour. Responses to all items (with the exception of pro-social behaviour items) are scored and totalled to generate the *Total Difficulties Score* (TDS), which can reach a maximum of 40 points. Presence of overall mental health problems (Exposure 1), was indicated by a TDS score of 17 or above, which is considered to indicate the “abnormal” mental health category (Goodman, 1997).

The emotional symptoms sub-scale assesses the presence of emotional problems (Exposure 2, including anxiety problems and low mood), and a score of 5 or above out of 10 indicates a high risk of emotional problems (Goodman, 1997). Importantly, this sub-scale has been shown to discriminate 7-11 year old children with and without diagnosable anxiety disorders within a community sample with a reasonable level of accuracy (Area Under the Curve >0.8, sensitivity, 75%, specificity 73%) (Reardon et al., 2025).

We conducted separate analyses for exposures related to overall mental health problems and emotional problems. A mental health “shock” for overall mental health problems was identified by the first time a child’s TDS was greater or equal to 17, and a “shock” for emotional problems was the first time a child’s emotional symptoms score was greater or equal to 5. Using data from four MCS surveys (wave 3 to 6), we created the “shock” indicators for each age of onset (5, 7, 11, 14). A value of 1 was assigned if a shock occurred at that age, and a value of 0 if a shock did not occur at that age. This means that for each mental health exposure (mental health problems and emotional problems), a maximum of one indicator was assigned a value of 1 (indicating age of onset).

### 2.3 Outcome and Mediator Variables

The outcomes in our primary analyses were a set of variables obtained from the NPD about GCSE performance in the academic year 2016/17 when children were 16 years old. Before 2017, GCSE subjects were graded by letters, with A* being the highest and G the lowest. In 2017, a numerical grading scheme ranging from 1 (the lowest) to 9 (the highest) started to replace the old one. These primary outcomes included: the number of subjects taken (from 0 to 14); the number of subjects passed; achieving five or more subjects with a grade of A*-C, including English and Maths; and the average numerical score for all subjects, where scores ranged from 1 (the lowest) to 9 (the highest) by design.

To complement the primary analyses, we also examined, in secondary analyses, school absenteeism and the likelihood of studying at university, and also explored the potential mediating role of absenteeism in the relationship between mental health problems and educational outcomes.

School absenteeism data was also obtained from the NPD. In England, a school day is divided into two sessions (morning and afternoon). With a minimum of 190 school days per academic year, schools must provide at least 380 sessions annually. The Department for Education collects the total number of sessions provided by schools and the actual number of sessions attended by each student from schools. We used this information from the academic year 2006/07 to 2016/17 to calculate the rate of school absence by dividing the total number of missed sessions by the total number of possible sessions over these years. Rate of school absence was used as a regressor in secondary analyses that explored whether school absenteeism could be a mediator of the relationship between mental health problems and GSCE outcomes.

The perceived likelihood of attending university was separately reported by the children and their main responding parent in the seventh survey (age 17). Both were asked a similar question about how likely the child was to study at university. However, their answer was recorded differently: children provided a percentage estimate ranging from 0 to 100, whereas parents chose an answer from a categorical variable with the following categories: “very likely”, “fairly likely”, “not very likely”, and “not at all likely”. To present children’s and parents’ responses on a consistent scale, we converted children’s evaluation into four groups, using 25, 50 and 75 as cut-off values. Then, we modelled both categorical outcomes using an ordered logit specification.

As data sources for estimating potential productivity losses associated with mental health problems in childhood partly rely on the estimates produced in primary and secondary analyses, we report both those and other external sources of data in Table 4, with further in-depth information detailed in Appendix A2 and Appendix Table A6.

### 2.4 Control variables

Among children and young people, emotional problems are frequently comorbid with conduct problems (Angold et al., 1999). Therefore, in the main analyses related to emotional problems, we adjusted for the onset of conduct problem at ages 5, 7, 11 and 14, defined by a score of 4 or higher in the SDQ conduct problem subscale (Goodman, 1997).

Across all models, we additionally controlled for socio-demographic characteristics, including information on children (sex, ethnicity, birthweight, year and month of birth), mother (age of childbirth, education, and maternal depression) and family (number of siblings, number of household members and income at baseline). In the analysis where the outcome variable was the number of GCSEs passed, the number of GCSEs taken was used as an additional control variable.

## 3. Methods

### 3.1 Regression Analyses

We estimated the association between mental health problems in children and adolescents and GCSE performance using the following regression model:

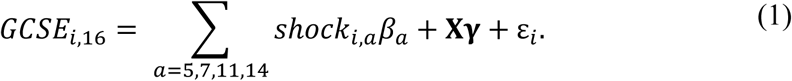

where 𝐺𝐶𝑆𝐸_𝑖,16_represents each of the four GCSE-related outcomes (i.e. number of subjects taken; number of subjects passed; achieving five or more subjects with a grade of A*-C, including English and Maths; and average numerical score of all subjects) of individual *i* at age 16 years. The main exposure in this analysis is 𝑠ℎ𝑜𝑐𝑘_𝑖,𝑎_, which captures the overall mental health shock (Exposure 1) or the emotional problems shock (Exposure 2) at age *a*, and 𝐗 represents a vector of control variables including socio-demographic characteristics for the child, the mother, and the family (see Section 2.4 for details). When we analysed the number of GCSEs passed, we additionally controlled for the number of GCSEs taken. As achieving five or more subjects with A*–C including English and Math is a binary outcome, we used a Probit model to estimate this outcome. Average marginal effects derived from the Probit model were reported instead of the original Probit model estimates, as the latter is for the latent variable that does not have a natural interpretation. We used linear regression for analysing the remaining GCSE outcomes (i.e. number of subjects taken; number of subjects passed; and average numerical score of all subjects).

We subsequently examined whether school attendance could explain the association between mental health shocks and GCSE outcomes. For doing that, we first considered school absenteeism as an outcome and regressed the school absence rate on the mental health shock indicators across ages. We then re-estimate Equation (1) by additionally including the absence rate for each different GCSE outcome.

Finally, as GCSE performance plays a critical role in university admission in the UK (Crawford, 2014), we replaced the GCSE outcomes in Equation (1) with perceived likelihood of higher education at 17 years to examine the association with mental health shocks in childhood and adolescence. We estimated an ordered logit model for these categorical responses.

All analyses were undertaken in STATA 18. The STATA survey commands “svy:” were used to account for the clustered, stratified design of the MCS with the provided non-response weights.

### 3.2 Estimating Productivity Loss

We also provided a simple model to estimate the potential productivity loss at both the individual and societal levels due to not achieving the required GCSE qualification (i.e. five or more GCSEs at grades A*–C/9–4), which is attributable to mental health problems and emotional problems between 5 and 14 years.

At the individual level, the expected loss of future income, ℓ, can be expressed by

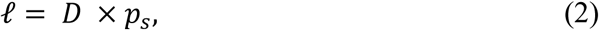

where 𝐷 is the labour market return on the GCSE qualification, which is captured by the lifetime earning difference between individuals achieving the GCSE qualification or not; 𝑝_𝑠_is the increased probability of failing to achieve GCSE qualification for a child experiencing a shock *s* at 5-14 years. In this study, a shock *s* represents a shock for either overall mental health problems (*Exposure 1*) or for emotional problems (*Exposure 2*) (i.e. 𝑠 ∈ {𝑒𝑥𝑝𝑜1, 𝑒𝑥𝑝𝑜2})

The probability 𝑝 can be further decomposed into

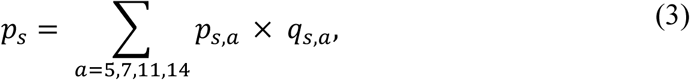

where 𝑝_𝑠,𝑎_denotes the probability of having a shock with the onset age *a* conditional on ever having such a shock at 5-14 years; 𝑞_𝑎_represents the probability of failing GCSE conditional on having this shock at age *a*. We obtained 𝑝_𝑠,𝑎_ from the descriptive statistics and 𝑞_𝑠,𝑎_ from the estimates obtained in Eq. (1) when the outcome was achieving five or more subjects with A*-C (including English and Maths) or not.

With the knowledge of individual productivity loss, the productivity loss in England, ℒ, can be calculated as follows:

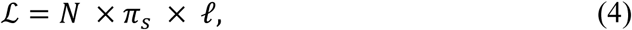

where 𝑁 is the population size of 14-year-olds in England in 2015, which is the total population in the same cohort of the MCS sample; 𝜋_𝑠_ is the probability that a child ever experiences a shock *s* at 5-14 years. For this calculation, we assumed that all probabilities in Equations (2)-(4) (i.e. 𝑝_𝑠,𝑎_, 𝑞_𝑠,𝑎_ and 𝜋_𝑠_), as derived from the MCS sample, also applied to children in the same cohort in England. The specific value of each parameter and its source are summarised in Table 4. Details on calculating the earning difference are provided in Appendix A2.

## 4. Results

### 4.1 Descriptive Statistics

The working sample for the present study consisted of 4,783 children and their families, and Appendix Figure A1 reports a flowchart of the inclusion/exclusion criteria of the sample. For analyses of the binary outcome ‘achieving five or more subjects with a grade of A*-C, including English and Maths’, we excluded a further 298 children who took fewer than five subjects.

Table 1 reports the weighted mean and standard deviation for the working sample. The children in our sample were born between 2000 and 2002, and their mothers had a mean age of 28.7 years at the time of birth. Nearly 90% of children were white and about half were male and half female. The average household size was 3.1 members, including 1.2 siblings. In the Academic year 2016/17, children took a mean of 9.1 GCSE subjects and passed 6.7 of them. Around 57% of the children achieved 5 GCSEs at grades A*–C including English and Maths. In terms of the numerical score, the average was around 4.7, which falls between a standard pass (4) and a strong pass (5).

**Table 1.**
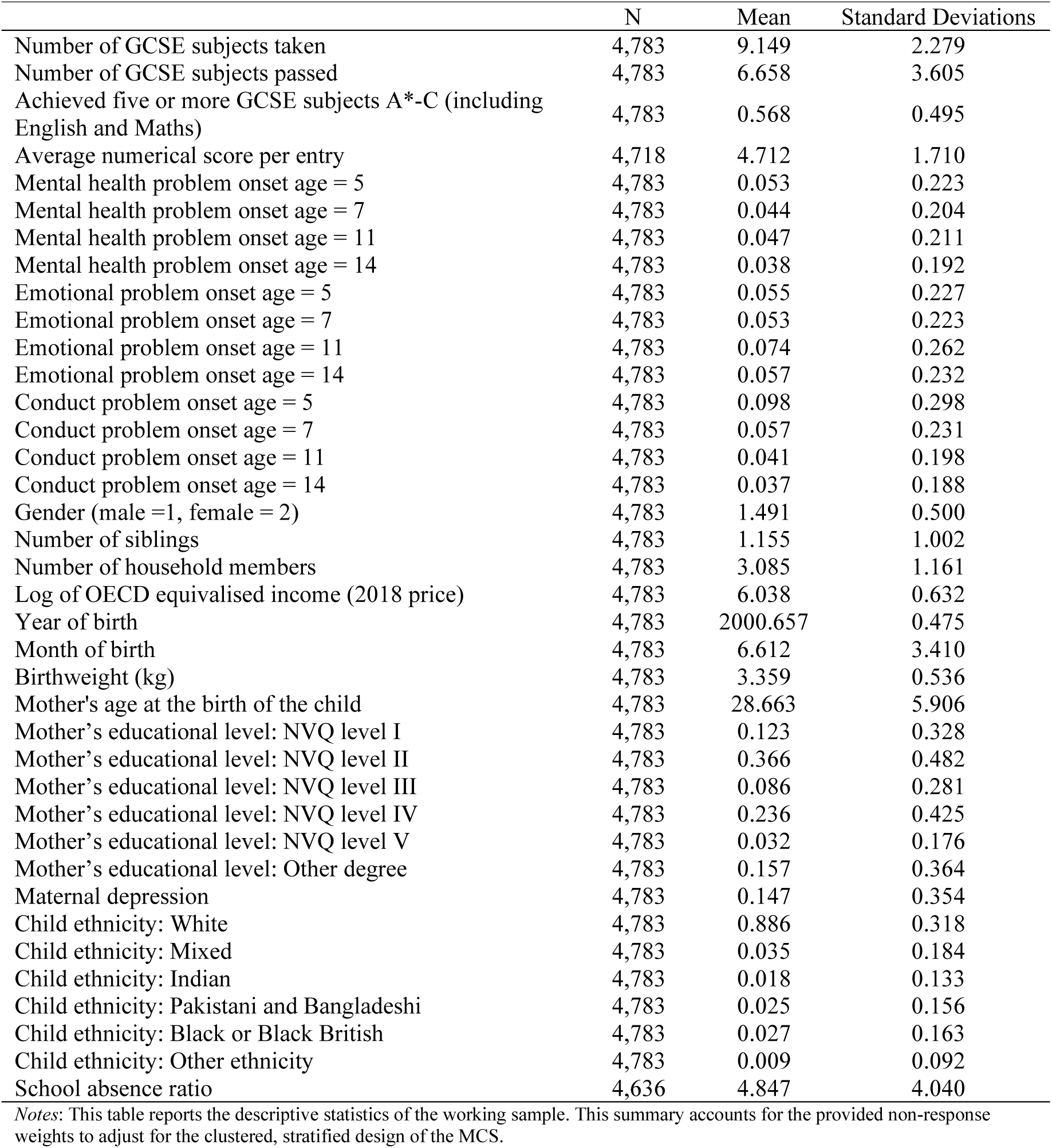
Descriptive Statistics.

Figure 1 summarises the percentages of health shocks across ages within our sample. The bars on the left and right represent the shocks measured by the SDQ TDS (overall mental health problems – Exposure 1) and the SDQ Emotional Symptoms Sub-scale (emotional problems – Exposure 2), respectively. In both charts, we found around 80% of children in our sample (specifically, 81.86% for Exposure 1, and 76.17% for Exposure 2) had never experienced any mental health shocks by age 14 years. However, the percentage of participants who had an emotional problem shock during this period (23.83%) was higher than that for overall mental problems (18.14%). Mental health shocks measured by the SDQ TDS were slightly more common at younger ages. In contrast, the probability of having an emotional problem shock was highest at age 11, and similar probabilities applied to other ages.

**Figure 1:**
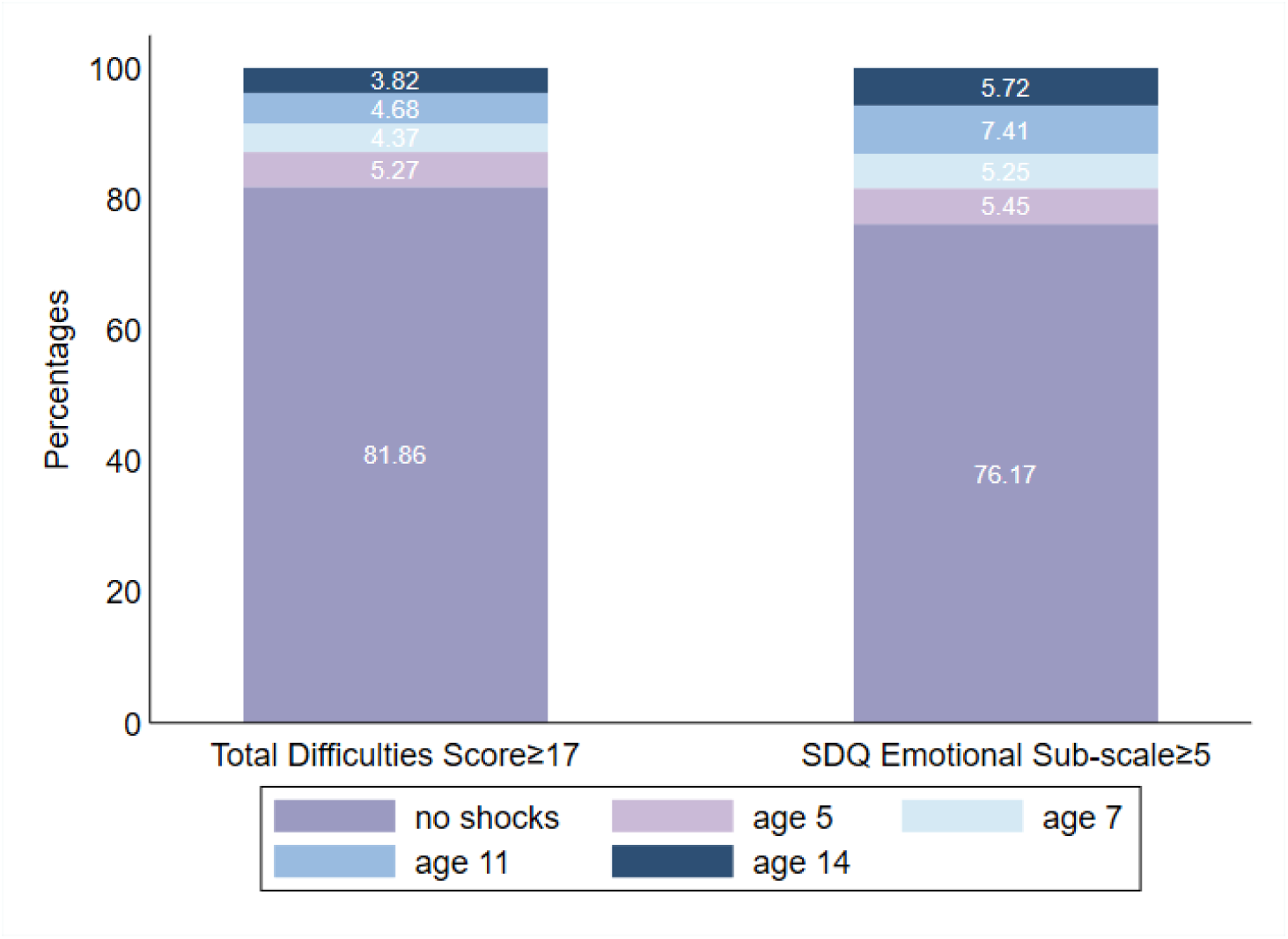
Mental health shocks across ages *Note*: This figure summarises the distribution of ages for mental health shocks of the working sample (N=4,783). The original summary can also be found in Table 1. The bar on the left is for the general mental disorders measured by the *Total Difficulty Score* (≥17); the bar on the right is for anxiety disorders measured by the *SDQ Emotional Symptoms Sub-scale* (≥5).

### 4.2 Mental health problems and educational achievement

Table 2 presents the association between the mental health problems (SDQ TDS) shock and GCSE performance. These shocks, at all age points, were negatively associated with all GCSE outcomes at the 1% significance level. Results showed that students who experienced a shock at any of the timepoints took on average 1.14-2.01 fewer GCSE subjects and passed 0.73-1.31 fewer GCSE subjects than those who did not. The chance of achieving five or more GCSEs at grades A*-C including English and Math was 18.6 to 24.9 percentage points lower for the group with than without a shock. With respect to GCSE numerical scores, a shock was correlated with an average reduction of 0.74-1.1 per GCSE subject. These negative associations were particularly strong at age 11 years, with the exception of the outcome ‘number of GCSEs taken’ where the largest negative association occurred at age 7 years.

**Table 2.** Association between Mental Health Shocks (Measured by the Total Difficulty Score≥17) and GCSE Performance.

|  | (1)<br>Number of GCSE<br>Subjects Taken | (2)<br>Number of GCSE<br>Subjects Passed | (3)<br>Five or More<br>Subjects with A*-<br>C, including<br>English and Math | (4)<br>Average Numerical<br>Score |
| --- | --- | --- | --- | --- |
| *General mental health problems* |  |  |  |  |
| Shock at Age 5 | -1.246***<br>(0.271) | -0.939***<br>(0.229) | -0.202***<br>(0.038) | -0.721***<br>(0.114) |
| Shock at Age 7 | -2.010***<br>(0.403) | -0.729***<br>(0.241) | -0.186***<br>(0.041) | -0.855***<br>(0.123) |
| Shock at Age 11 | -1.379***<br>(0.230) | -1.311***<br>(0.208) | -0.249***<br>(0.039) | -1.101***<br>(0.110) |
| Shock at Age 14 | -1.135***<br>(0.253) | -0.952***<br>(0.231) | -0.216***<br>(0.041) | -0.856***<br>(0.136) |
| Observations | 4,783 | 4,783 | 4,491 | 4,718 |

*Notes:* This table reports the associations between mental health shocks measured by the Total Difficulty Score (TDS ≥ 17) and GCSE outcomes. Models presented in Columns (1)-(4) all control for children's sex, ethnicity, birthweight, year and month of birth, mother's age of birth, education, maternal depression, number of siblings, number of household members and income at the baseline. Column (2) additionally controls for the number of GCSE subjects taken. Column (3) restricts the sample to children taking more than five GCSE subjects. It estimates a Probit model and presents the average marginal effects of the corresponding estimates. Column (4) restricts the sample to children with available numerical GCSE scores. Statistical significance levels are shown as \*\*\* p<.01, \*\* p<.05, \* p<.1.

Table 3 reports the analyses focusing on the mental health shocks resulting from emotional problems. Although emotional problems shocks at each age were negatively associated with all GCSE outcomes, only some of these associations were statistically significant in the main analysis where we controlled for the onset of conduct problems. In these analyses, notably, onset of emotional problems at age 11 was significantly associated with each GCSE outcome, including a roughly 14 percentage point reduction in the chance of achieving 5 or more GCSEs at grade A*-C including Maths and English. Moreover, onset of emotional problems at age 5 showed a very similar statistically significant association with achieving these 5 GCSEs. Appendix Table 1 presents the results for the emotional problems shocks without adjusting for conduct problems. It is notable that these associations are larger in magnitude than those presented in Table 3, and with one exception, all were statistically significant at the 5% level.

**Table 3.** Association between Mental Health Shocks (Measured by the SDQ Emotional Symptoms Sub- cale≥5) and GCSE Performance.

|  | (1)<br>Number of GCSE<br>Subjects Taken | (2)<br>Number of GCSE<br>Subjects Passed | (3)<br>Five or More<br>Subjects with A*-<br>C, including<br>English and Math | (4)<br>Average Numerical<br>Score |
| --- | --- | --- | --- | --- |
| *Emotional problems* |  |  |  |  |
| Shock at Age 5 | -0.152<br>(0.183) | -0.283<br>(0.233) | -0.134***<br>(0.037) | -0.148<br>(0.105) |
| Shock at Age 7 | -1.001***<br>(0.316) | 0.012<br>(0.219) | -0.047<br>(0.037) | -0.265**<br>(0.129) |
| Shock at Age 11 | -0.945***<br>(0.208) | -0.496***<br>(0.158) | -0.146***<br>(0.030) | -0.454***<br>(0.086) |
| Shock at Age 14 | -0.515***<br>(0.163) | -0.253<br>(0.200) | -0.063*<br>(0.037) | -0.412***<br>(0.095) |
| *Conduct Problems* |  |  |  |  |
| Shock at Age 5 | -0.789***<br>(0.167) | -0.864***<br>(0.165) | -0.142***<br>(0.030) | -0.619***<br>(0.080) |
| Shock at Age 7 | -1.185***<br>(0.306) | -0.584**<br>(0.263) | -0.135***<br>(0.040) | -0.630***<br>(0.126) |
| Shock at Age 11 | -0.790***<br>(0.235) | -0.795***<br>(0.272) | -0.081*<br>(0.044) | -0.657***<br>(0.129) |
| Shock at Age 14 | -0.900***<br>(0.215) | -0.951***<br>(0.240) | -0.178***<br>(0.043) | -0.698***<br>(0.134) |
| Observations | 4,783 | 4,783 | 4,491 | 4,718 |

*Notes:* This table reports the associations between mental health shocks measured by the SDQ Emotional Symptoms Sub-scale ( $\geq 5$ ) and GCSE outcomes. A set of health shock indicators of behavioural problems measured by the SDQ conduct problems sub-scale ( $\geq 4$ ) is controlled for as common comorbidity. Models presented in Columns (1)-(4) all control for children's sex, ethnicity, birthweight, year and month of birth, mother's age of birth, education, maternal depression, number of siblings, number of household members and income at the baseline. Column (2) additionally controls for the number of GCSE subjects taken. Column (3) restricts the children to those taking more than five GCSE subjects. It estimates a Probit model and presents the average marginal effects of the corresponding estimates. Column (4) restricts the sample to children with available numerical GCSE scores. Statistical significance levels are shown as \*\*\* $p < .01$ , \*\* $p < .05$ , \* $p < .1$ .

**Table 4.** Model Parameters to Calculate Productivity Loss.

| Equation | Parameter | Description | Value | Source |
| --- | --- | --- | --- | --- |
| 2 | $D$ | Earning difference between people with and without the GCSE qualification | £109,250.35 | Details available in Appendix A2 |
| 3 | $p_{expo1,a}$ | The probability of the onset age = $a$ given having mental health problems between 5-14 years | $p_{mh,5} = 29.0\%$,<br>$p_{mh,7} = 24.1\%$,<br>$p_{mh,11} = 25.8\%$,<br>$p_{mh,14} = 21.1\%$ | Rescaled using the probabilities reported in Table 1 |
| 3 | $q_{expo1,a}$ | Probability of not achieving GCSE given the mental health problem onset age = $a$ | $q_{mh,5} = 20.2\%$,<br>$q_{mh,7} = 18.6\%$,<br>$q_{mh,11} = 24.9\%$,<br>$q_{mh,14} = 21.6\%$ | Regression Analysis (Table 2, Column 3) |
| 3 | $p_{expo2,a}$ | The probability of the onset age = $a$ given having emotional problems between 5-14 years | $p_{emo,5} = 22.9\%$,<br>$p_{emo,7} = 22.0\%$,<br>$p_{emo,11} = 31.1\%$,<br>$p_{emo,14} = 24.0\%$ | Rescaled using the probabilities reported in Table 1 |
| 3 | $q_{expo2,a}$ | Probability of not achieving GCSE given the emotional problem onset age = $a$ | $q_{emo,5} = 13.4\%$,<br>$q_{emo,7} = 4.7\%$,<br>$q_{emo,11} = 14.6\%$,<br>$q_{emo,14} = 6.3\%$ | Regression Analysis (Table 3, Column 3) |
| 4 | $N$ | The population size of the MCS cohort in England | 608,240 | Official Statistics: Revised population estimates for England and Wales: mid-2012 to mid-2016 (Sheet “MYE – 2014 All”) |
| 4 | $\pi_{expo1}$ | The probability of having mental health problems between 5-14 years | 18.14% | Derived from the probabilities reported in Table 1 |
| 4 | $\pi_{expo2}$ | The probability of having emotional problems between 5-14 years | 23.83% | Derived from the probabilities reported in Table 1 |

### 4.3 Mediating Role of School Absence

Overall, the absence rate was around 4.8% throughout these academic years (see Table 1). Appendix Table A2 reports on the results of regressing the school absence rate on the mental health shock indicators across ages. All mental health shock indicators, regardless of age and measurement, were correlated with a higher absence rate. Appendix Tables A3 and A4 present results from re-estimating Equation (1), for each GCSE outcome, when the absence rate was included as an additional regressors. Our findings suggest that school absence was strongly and negatively associated with GCSE performance. A one-standard-deviation increase in absence rate (4.04 percentage points) was associated with a reduction in 0.76 GCSE subjects taken, 0.27 GCSE subjects passed, a decline in the probability of achieving five or more GCSE subjects at the grade of A*-C by 10 percentage points, and the average numerical scores by 0.37 points.

To explore the potential mediating effect of school absence, we compared the estimates of mental health shocks from Appendix Tables A3 and A4 to those from the main models without controlling for school absence (shown in Tables 2 and 3). For ease of comparison, we re-estimated the main model based on the same sample of this mediating analysis and plotted the estimates from both regression in Appendix Figures A2.1 and A2.2.

Results in both figures suggested partial mediating effects of school absence, as the negative association between mental health shocks and GCSE outcomes diminished in magnitude when the school absence rate was controlled for. For mental health shocks based on the SDQ TDS (Appendix Figures A2.1), the mediating effect was particularly strong for the outcome “number of GCSE subjects taken”, where including school absence led to a 24%-34% reduction in the health shock estimates for different onset ages. We also found moderate mediating effects for other GCSE outcomes with reductions of around 10%-20%. For shocks based on the SDQ Emotional Symptoms Sub-scale (Appendix Figures A2.2), the mediating effect was larger in magnitude, except for the outcome “number of GCSE subjects passed”. For the other GCSE-related outcomes, school absence explained at least 30% of the negative association for most indicators at different onset ages.

### 4.4 Mental Health Shocks and Likelihood of Higher Education

Appendix Table A5 reports the association between mental health shocks and the perceived likelihood of entering university, where the estimates are expressed in terms of odds ratios. An odds ratio greater than one means a higher possibility of achieving high-ranking outcomes than lower-ranking outcomes while a ratio smaller than one implies a lower possibility.

Results consistently indicated that both children and their parents were less optimistic about studying at the University when the child had experienced a mental health or emotional problems shock during childhood and adolescence studying. These findings also highlighted that ages 7 and 11 years as particularly critical periods in which a mental health shock had a stronger negative association based on both child and parent report. This was especially evident when the mental health shocks were measured by the TDS, and the outcome was parent-reported, with the highest likelihood found at age 14 years.

### 4.5 Potential Productivity Loss

The value (2023 price) of the labour market return to GCSE between ages 23 and 68 years was estimated at £109,250.35 per person, based on a wage premium of 1.25 for individuals with the GCSE qualification (Mcintosh, 2006) and the minimum wage rate England in 2022 (GOV.UK, 2024a). The increased probability of not achieving the GCSE qualification associated with mental health problems and emotional problems between ages 5 and 14 was 21.32% and 10.16%, respectively. Hence, the expected earning loss was £23,294 for mental health problems and £11,111 for emotional problems at the individual level. According to Equation (4), the corresponding productivity loss in England was estimated at approximately £2.57 billion for mental health problems and £1.61 billion for emotional problems for the entire population born in the same period as the MCS cohort (i.e. 2000/02).

## 6. Discussion

In this study, we found that, between ages 5 and 14 years, significant groups of children and adolescents in England experienced an emotional problem shock (23.83%) or an overall mental health problem shock (18.14%). Overall mental health problem shocks were likely to be somewhat less common because they were identified using the SDQ TDS, a broader measure of emotional and behavioural symptoms with a higher threshold for classification in the “abnormal” range than the emotional problem scale.

More importantly, this study highlighted the negative association between early onset of mental health problems in children and adolescents and educational attainment at age 16. Onsets of mental health problems at age 5, 7, 11 and 14 were each associated with poor GCSE outcomes, including the number of subjects taken, the number of subjects passed, the possibility of achieving five or more subjects at grade of A*-C (including English and Maths), and numerical GCSE grades. Statistically significant negative associations were also found for emotional problems that onset at age 5, 7, 11 and 14 and some GCSE outcomes.

However, notably, it was for emotional problems that first occurred at age 11 where results were most stark, with statistically significant negative associations across all GCSE outcomes, even after adjusting for co-occurring conduct problems. Age 11 coincides with transition from primary to secondary school education and has been previously highlighted as a vulnerable period for mental health (Donaldson et al., 2024; Rice et al., 2011).

This study also provided evidence to suggest that a considerable part of the negative association can be explained by school absenteeism. Our findings replicated the established link between mental health problems and school absenteeism (e.g., Finning et al., 2020; Lester & Michelson, 2024), but extended this by demonstrating that absenteeism accounted for more than one-quarter of the negative association with the “number of GCSE subjects taken”. The mediating effect was also strong for the association between emotional problem shock and “GCSE grades”. These findings support recent calls to improve school attendance and educational outcomes through early, effective intervention for child and adolescent mental health problems (GOV.UK, 2024b).

We also found evidence that childhood and adolescent mental health shocks are associated with lower perceived likelihood of higher education, as reported by both children and their parents. The strongest associations were concentrated around age 11, again emphasising this age as a salient period for subsequent educational trajectories. These findings complement existing evidence linking mental health-related symptoms to aspirations. For instance, a large population-based study of Norwegian adolescents reported that higher levels of social anxiety between ages 13 and 15 years, assessed using both diagnostic interviews and self-reported measures, were associated with reduced likelihood of aspiring to higher education, with odds ratios of 0.74 for adolescents screening positive on the ADIS-C (Anxiety Disorders Interview Schedule for Children), and 0.92 for each unit increase in the SPAI-C score (Social Phobia and Anxiety Inventory for Children) (Jystad et al., 2021).

We estimated the potential labour income and productivity loss associated with overall mental health problems (Exposure 1) and emotional problems (Exposure 2) during childhood and early adolescence to be over £23,000 and £11,000 per affected child, respectively. This corresponds to potential productivity losses of £2.57 billion and £1.6 billion, respectively, within the population of England who were born during the same period as the MCS cohort (i.e. 2000/02). To provide a sense of magnitude, these losses represents about 18% and 11% of the NHS budget for mental health, learning disability and dementia services in 2023/24 (£14.4 billion) (Baker & Kirk-Wade, 2024). The number will be massively scaled up when more cohorts, especially younger cohorts, are taken into account. This result provides further justification for policy to drive effective prevention and early intervention for youth mental health problems, and particularly in the primary school age group.

Strengths of this study include a model specification that allows a comparison between the strength of the association between onset of mental health problems and educational outcomes across four different onset ages spanning 5-14 years. An alternative model specification could have used four non-mutually exclusive indicators to capture the presence of mental health difficulties at each survey, rather than focusing on onset. However, our method provides the overall association between the early onset of mental health difficulties and GCSE performance, which accounts for the possibility that mental health problems may persist over time, and examines the heterogeneity in the association of mental health shocks and GCSE performance across onset ages. This approach facilitated a comparison between the coefficients on mental health shocks at ages 5, 7, 11 and 14 years, highlighting a particularly strong association at age 11, which is meaningful in terms of timing of prevention and early intervention.

A further strength is the provision of a framework to estimate the potential productivity loss to an individual and society related to the early onset of overall mental health problems and emotional problems in particular. Health at young ages has been recognised as important human capital that entails more educational opportunities and higher future income (Currie, 2020). However, the evidence on human capital loss in relation to overall mental health problems and emotional problems specifically in childhood is still limited. The substantial productivity losses highlighted by this study underpin the need for ongoing initiatives to increase access to mental health services for children and young people.

This study also has some limitations. First, our results do not allow for causal interpretation. Despite a variety of covariates having been controlled for, unobserved confounding variables may still exist, which may raise an omitted variable bias. Furthermore, despite parents being the recommended reporters to enable reliable measurement across our age ranges (Bergström & Baviskar, 2021), the identification of a causal relationship may also be undermined by measurement error due to use of the parent-reported SDQ. Additionally, our analytic sample is predominantly white (approximately 90%), which may limit the representativeness of other ethnic groups and the generalisability of the findings. In addition, our calculation of productivity loss relied solely on the wage premium of obtaining five or more GCSE subjects with A*-C including English and Maths. This approach neglects the marginal labour market return between grades within A*-C (e.g., A* vs. A or B vs. C) and the heterogeneous return among subjects (Hodge et al., 2021). Moreover, the estimation naïvely assumes similar labour market participation between the high- and low-education groups. However, in the UK, workers with low education are more likely to have involuntary part-time jobs or to leave the labour market (OECD, 2024). Therefore, our calculation should be considered as a lower bound, which excludes other potential channels that could contribute to a greater labour income difference.

Despite these limitations, this study highlights the negative association between mental health problems during childhood and adolescence and educational outcomes. The related long-term earning and productivity loss point to the likely value of implementing collaborative strategies to support students with mental health problems to enhance their academic performance and future prospects.

## CRediT authorship contribution statement

**Shuye Yu**: Conceptualization, Methodology, Software, Formal analysis, Data Curation, Writing – Original Draft, Writing – Review & Editing Draft, Visualization. **Jack Pollard**: Conceptualization, Methodology, Formal analysis, Data Curation, Writing – Review & Editing Draft. **Tessa Reardon**: Conceptualization, Methodology, Writing – Review & Editing Draft. **Cathy Creswell**: Conceptualization, Methodology, Writing – Review & Editing Draft, Funding Acquisition. **Ruth Wadman:** Conceptualization, Methodology, Writing – Review & Editing Draft. **Mara Violato**: Conceptualization, Methodology, Supervision, Project Administration, Writing – Original Draft, Writing – Review & Editing Draft, Funding Acquisition.

## Ethics approval

The Millennium Cohort Study was granted ethical approval by multiple National Health Service Research Ethics Committees. No additional ethical approval was required for this study secondary analyses.

## Acknowledgements

We would like to express our gratitude to all the families who took part in the Millennium Cohort Study, without whom this research would not have been possible. We also thank the Centre for Longitudinal Study team, University of London, for producing the relevant data documentation, and the UK Data Service for making the data available. Finally, we are grateful to the Institutions that funded this work.

## Data availability

The authors do not have permission to share data, but data from the UK Millennium Cohort Study is openly available on the UK Data Service (https://ukdataservice.ac.uk/).

## References

Anderson, O. (2023). Walking the line: Does crossing a high stakes exam threshold matter for labour market outcomes? International Journal of Population Data Science, 8(2), Article 2. 10.23889/ijpds.v8i2.2187

Angold, A., Costello, E. J., & Erkanli, A. (1999). Comorbidity. Journal of Child Psychology and Psychiatry, 40(1), 57–87. 10.1111/1469-7610.00424

Baker, C., & Kirk-Wade, E. (2024). Mental health statistics: Prevalence, services and funding in England. https://commonslibrary.parliament.uk/research-briefings/sn06988/

Benton, T. D., Boyd, R. C., & Njoroge, W. F. M. (2021). Addressing the Global Crisis of Child and Adolescent Mental Health. JAMA Pediatrics, 175(11), 1108–1110. 10.1001/jamapediatrics.2021.2479

Bergström, M., & Baviskar, S. (2021). A Systematic Review of Some Reliability and Validity Issues regarding the Strengths and Difficulties Questionnaire Focusing on Its Use in Out-of-Home Care. Journal of Evidence-Based Social Work, 18(1), 1–31. 10.1080/26408066.2020.1788477

Butterworth, P., & Leach, L. S. (2018). Early Onset of Distress Disorders and High-School Dropout: Prospective Evidence From a National Cohort of Australian Adolescents. American Journal of Epidemiology, 187(6), 1192–1198. 10.1093/aje/kwx353

Card, D. (1999). The Causal Effect of Education on Earnings. In O. C. Ashenfelter & D. Card (Eds), Handbook of Labor Economics (Vol. 3, pp. 1801–1863). Elsevier. 10.1016/S1573-4463(99)03011-4

Cornaglia, F., Crivellaro, E., & McNally, S. (2015). Mental health and education decisions. Labour Economics, 33, 1–12. 10.1016/j.labeco.2015.01.005

Crawford, C. (2014). The link between secondary school characteristics and university participation and outcomes. The IFS. 10.1920/re.ifs.2024.0631

Currie, J. (2020). Child health as human capital. Health Economics, 29(4), 452–463. 10.1002/hec.3995

Currie, J., Stabile, M., & Jones, L. (2014). Do stimulant medications improve educational and behavioral outcomes for children with ADHD? Journal of Health Economics, 37, 58– 69. 10.1016/j.jhealeco.2014.05.002

Dalsgaard, S., McGrath, J., Østergaard, S. D., Wray, N. R., Pedersen, C. B., Mortensen, P. B., & Petersen, L. (2020). Association of Mental Disorder in Childhood and Adolescence With Subsequent Educational Achievement. JAMA Psychiatry, 77(8), 797–805. 10.1001/jamapsychiatry.2020.0217

Dona, S. W. A., McKenna, K., Ho, T. Q. A., Bohingamu Mudiyanselage, S., Seymour, M., Le, H. N. D., & Gold, L. (2025). Health-related quality of life, service utilisation and costs for anxiety disorders in children and young people: A systematic review and meta-analysis. Social Science & Medicine, 373, 118023. 10.1016/j.socscimed.2025.118023

Donaldson, C., Hawkins, J., Rice, F., & Moore, G. (2024). Trajectories of mental health across the primary to secondary school transition. *JCPP Advances*, e12244. 10.1002/jcv2.12244

Finning, K., Ford, T., Moore, D. A., & Ukoumunne, O. C. (2020). Emotional disorder and absence from school: Findings from the 2004 British Child and Adolescent Mental Health Survey. European Child & Adolescent Psychiatry, 29(2), 187–198. 10.1007/s00787-019-01342-4

Finning, K., Ukoumunne, O. C., Ford, T., Danielson-Waters, E., Shaw, L., Romero De Jager, I., Stentiford, L., & Moore, D. A. (2019). Review: The association between anxiety and poor attendance at school – a systematic review. Child and Adolescent Mental Health, 24(3), 205–216. 10.1111/camh.12322

Goodman, R. (1997). The Strengths and Difficulties Questionnaire: A Research Note. Journal of Child Psychology and Psychiatry, 38(5), 581–586. 10.1111/j.1469-7610.1997.tb01545.x

Goodman, R. (1999). The Extended Version of the Strengths and Difficulties Questionnaire as a Guide to Child Psychiatric Caseness and Consequent Burden. The Journal of Child Psychology and Psychiatry and Allied Disciplines, 40(5), 791–799. 10.1111/1469-7610.00494

Goodman, R. (2001). Psychometric Properties of the Strengths and Difficulties Questionnaire. Journal of the American Academy of Child & Adolescent Psychiatry, 40(11), 1337–1345. 10.1097/00004583-200111000-00015

GOV.UK. (2024a). National Minimum Wage and National Living Wage rates. https://www.gov.uk/national-minimum-wage-rates

GOV.UK. (2024b). Working together to improve school attendance. https://www.gov.uk/government/publications/working-together-to-improve-school-attendance

Hansen, K. (2014). Millennium Cohort Study: A Guide to the Datasets (Eighth Edition).

Institute of Education University of London. https://cls.ucl.ac.uk/wp-content/uploads/2017/07/MCS-Guide-to-the-Datasets-022014.pdf

Hodge, L., Little, A., & Weldon, M. (2021). GCSE attainment and lifetime earnings. Department for Education. https://www.gov.uk/government/publications/gcse-attainment-and-lifetime-earnings

Jerrim, J. (2023). The benefits of meeting key grade thresholds in high-stakes examinations. New evidence from england. British Journal of Educational Studies, 71(1), 5–28. 10.1080/00071005.2022.2033692

Jystad, I., Haugan, T., Bjerkeset, O., Sund, E. R., & Vaag, J. (2021). School Functioning and Educational Aspirations in Adolescents With Social Anxiety—The Young-HUNT3 Study, Norway. Frontiers in Psychology, 12. 10.3389/fpsyg.2021.727529

Kieling, C., Buchweitz, C., Caye, A., Silvani, J., Ameis, S. H., Brunoni, A. R., Cost, K. T., Courtney, D. B., Georgiades, K., Merikangas, K. R., Henderson, J. L., Polanczyk, G. V., Rohde, L. A., Salum, G. A., & Szatmari, P. (2024). Worldwide Prevalence and Disability From Mental Disorders Across Childhood and Adolescence: Evidence From the Global Burden of Disease Study. JAMA Psychiatry, 81(4), 347–356. 10.1001/jamapsychiatry.2023.5051

Lester, K. J., & Michelson, D. (2024). Perfect storm: Emotionally based school avoidance in the post-COVID-19 pandemic context. BMJ Ment Health, 27(1). 10.1136/bmjment-2023-300944

Mcintosh, S. (2006). Further Analysis of the Returns to Academic and Vocational Qualifications. Oxford Bulletin of Economics and Statistics, 68(2), 225–251. 10.1111/j.1468-0084.2006.00160.x

Moore, G., Anthony, R. E., Hawkins, J., Van Godwin, J., Murphy, S., Hewitt, G., & Melendez-Torres, G. J. (2020). Socioeconomic status, mental wellbeing and transition to secondary school: Analysis of the School Health Research Network/Health Behaviour in School-aged Children survey in Wales. British Educational Research Journal, 46(5), 1111–1130. 10.1002/berj.3616

NHS England. (2023). Mental Health of Children and Young People in England, 2023—Wave 4 follow up to the 2017 survey. NHS England Digital. https://digital.nhs.uk/data-and-information/publications/statistical/mental-health-of-children-and-young-people-in-england/2023-wave-4-follow-up

OECD. (2024). How does educational attainment affect participation in the labour market? In Education at a Glance 2024 (pp. 78–96). OECD Publishing. 10.1787/f011522f-en

Polanczyk, G. V., Salum, G. A., Sugaya, L. S., Caye, A., & Rohde, L. A. (2015). Annual Research Review: A meta-analysis of the worldwide prevalence of mental disorders in children and adolescents. Journal of Child Psychology and Psychiatry, 56(3), 345–365. 10.1111/jcpp.12381

Pollard, J., Reardon, T., Williams, C., Creswell, C., Ford, T., Gary, A., Roberts, N., Stallard, P., Ukoumunne, O., & Violato, M. (2023). The multifaceted consequences and economic costs of child anxiety problems: A systematic review and meta-analysis. JCPP Advances, 3(3), e12149. 10.1002/jcv2.12149

Reardon, T., Ukoumunne, O. C., Ball, S., Brown, P., Ford, T., Gray, A., Hill, C., Jasper, B., Larkin, M., Macdonald, I., Morgan, F., Sancho, M., Sniehotta, F. F., Spence, S. H., Stainer, J., Stallard, P., Violato, M., Team, iCATS, & Creswell, C. (2025). Development of a brief assessment tool to identify children with probable anxiety disorders. JCPP Advances, 5(2), e12265. 10.1002/jcv2.12265

Rice, F., Frederickson, N., & Seymour, J. (2011). Assessing pupil concerns about transition to secondary school. British Journal of Educational Psychology, 81(2), 244–263. 10.1348/000709910X519333

Solmi, M., Radua, J., Olivola, M., Croce, E., Soardo, L., Salazar de Pablo, G., Il Shin, J., Kirkbride, J. B., Jones, P., Kim, J. H., Kim, J. Y., Carvalho, A. F., Seeman, M. V., Correll, C. U., & Fusar-Poli, P. (2022). Age at onset of mental disorders worldwide: Large-scale meta-analysis of 192 epidemiological studies. Molecular Psychiatry, 27(1), 281–295. 10.1038/s41380-021-01161-7

University College London, UCL Institute of Education, Centre for Longitudinal Studies, & Department for Education. (2022). Millennium Cohort Study: Linked Education Administrative Datasets (National Pupil Database), England: Secure Access [Data set]. 10.5255/UKDA-SN-8481-2

van Poortvliet, M. (2024). Child mental health and educational attainment: Longitudinal evidence from the UK. SSM - Mental Health, 5, 100294. 10.1016/j.ssmmh.2023.100294

Zając, T., Perales, F., Tomaszewski, W., Xiang, N., & Zubrick, S. R. (2024). Student mental health and dropout from higher education: An analysis of Australian administrative data. Higher Education, 87(2), 325–343. 10.1007/s10734-023-01009-9

