## Appendix for "Early Onset Mental Health Problems, Educational Attainment and Productivity Loss in England: Evidence from the Millennium Cohort Study"

**Appendix A1. GCSE in England**

The national curriculum in England is divided into four key stages. At ages 14-16 years, students enter into Key Stage 4, including Years 10-11. During this period, students select a range of subjects to study, typically around eight to ten, with Maths, English, and Science being compulsory. At the end of this stage, students' educational attainment is assessed by an examination named the General Certificate of Secondary Education (GCSE).

Before 2017, GCSE subjects were graded by letters, with A* being the highest and G the lowest. In 2017, a numerical grading scheme ranging from 1 (the lowest) to 9 (the highest) started to replace the old one. While the GCSE subjects taken by the cohort children in this study were originally graded with letters, the database we use also provides the corresponding numerical grades, which enables us to analyse their GCSE performance under both grading schemes.

It is important for students to achieve five or more GCSEs at grades A*- C/9-4. This is not only a benchmark indicator reviewed by the Department for Education annually, but also the minimum entry requirement to progress into higher academic qualifications, such as A-level and university [1]. This criterion is also widely recognised as an essential qualification by UK employers in their recruitment processes.

Wales and Northern Ireland also adopt the GCSE qualification, but the grading schemes and subject contents are different [2]. Scotland follows different academic qualifications from the GCSE.

**Appendix A2. Labour Market Return to GCSE**

We assume most individuals finish their education at the age of 23, after which a proportion enters the labour market. While individuals with low educational attainment can be active in the labour market and receive earnings at an earlier age, we exclude this early income for ease of comparison. Due to a recent pension age reform in the UK, the MCS cohort will receive their state pension at the age of 68, which incentivises them to work until this age. Therefore, the present value of earnings over this period can be calculated as follows:

$$Discounted\ Earning\ = \sum_{age=23}^{68} \frac{wage\ \times (1+\beta)^{age-23}}{(1+\alpha)^{age-23}}, \tag{A1}$$

where *wage* is the annual labour income; $\beta$ is the annual growth rate of the labour income against inflation; $\alpha$ is the discount factor reflecting one's time preference.

Furthermore, we also account for the fact that individuals may participate in the labour market to a different extent. For example, someone may opt themselves out for family or health reasons. Alternatively, they may opt for a part-time job rather than a full-time job. Hence, the *wage* parameter in Equation (1) can be decomposed into

$$wage\ = p_{job} \times \left[p_f\ \times h_f\ \times rate + \left(1 - p_f\right) \times h_p\ \times rate\right] \times 52, \tag{A2}$$

where $p_{job}$ is the proportion of the working-age population active in the labour force; $p_f$ is the proportion of workers in full-time employment and part-time employment otherwise; $h_f$ and $h_p$ are the weekly working hours for a full-time and part-time job, respectively; $rate$ is the hourly rate. To convert weekly labour income into yearly one, we multiply the weekly working hours by 52.

The low-education group tends to receive lower labour market returns than the high-education group. In the context of the GCSE qualification, the difference is characterised by a wage premium of 1.25, as documented in the labour economics literature.[3] This means Equations (A1) and (A2), on their own, can represent the labour income of individuals without the GCSE qualification. To express the labour income of individuals with the GCSE qualification, we replace the $rate$ parameter in Equation (A2) with $1.25\ \times\ rate$. The values of all parameters in Equations (A1) and (A2) are summarised in Appendix Table A7 below.

We assume the minimum wage rate in 2023 (i.e. £10.42) applies to individuals without the GCSE qualification. This makes our calculation a conservative estimation of the labour income difference. In reality, the low-education group may earn above the minimum wage, which would enlarge the difference when we apply the wage premium of 1.25 to the high-education group.

By inserting all values reported in Appendix Table A7 into Equations (A1) and (A2), we find the difference in labour market return is £109,250.35 between individuals with and without a GCSE qualification over the ages of 23 to 68. This value serves as the value for parameter $D$ in Equation (2) in Section 6 of the main text.

**Appendix Figure A1. Flowchart of the Working Sample**

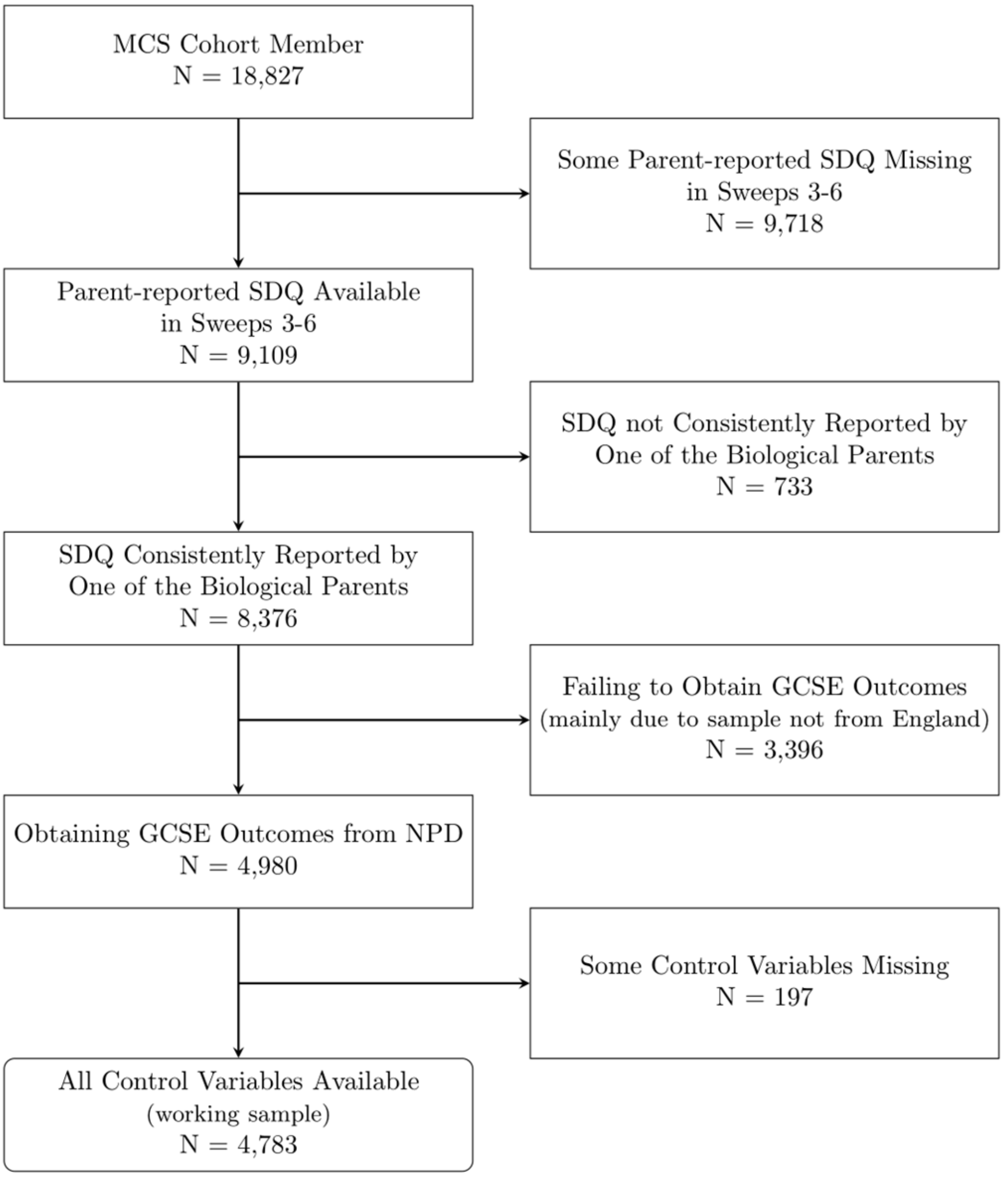

**Appendix Figure A2.1. Association between Mental Health Shocks (TDS≥17) and GCSE Outcomes**

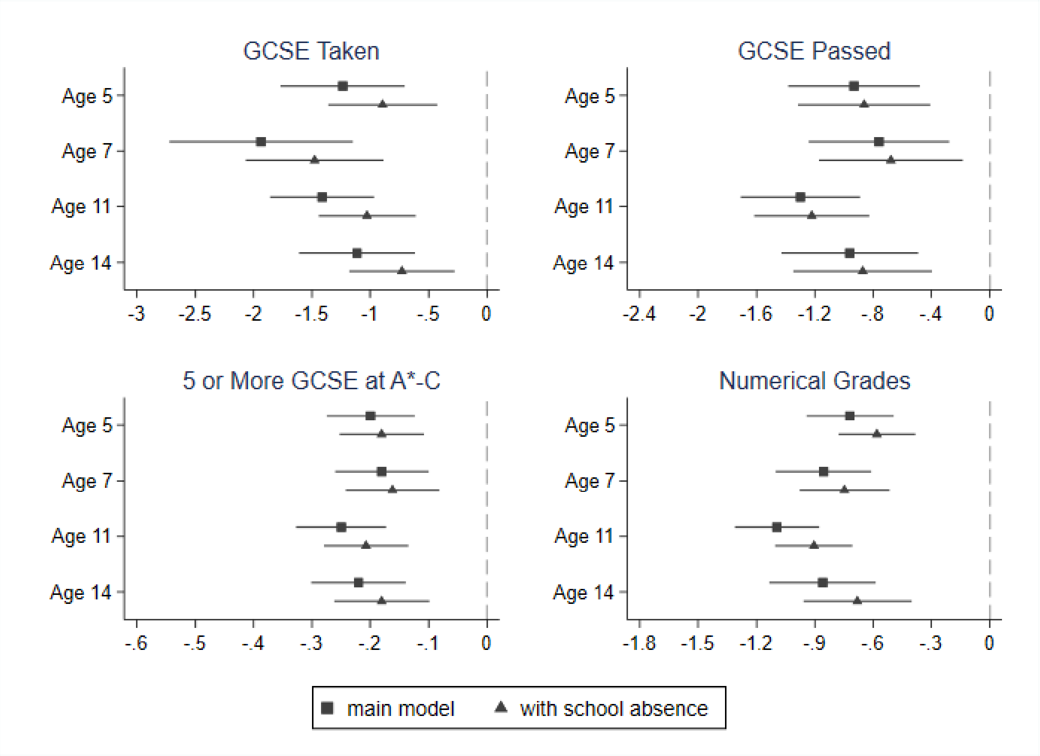

*Notes*: This figure plots the estimates from the main analysis (Table 2) and the one with the additional control of school absence (Appendix Table A3) based on the sample for whom the school attendance data are available (n=4,636).

**Appendix Figure A2.2. Association between Emotional Problem Shocks (SDQ Emotional Symptoms Sub-scale≥5) and GCSE Outcomes**

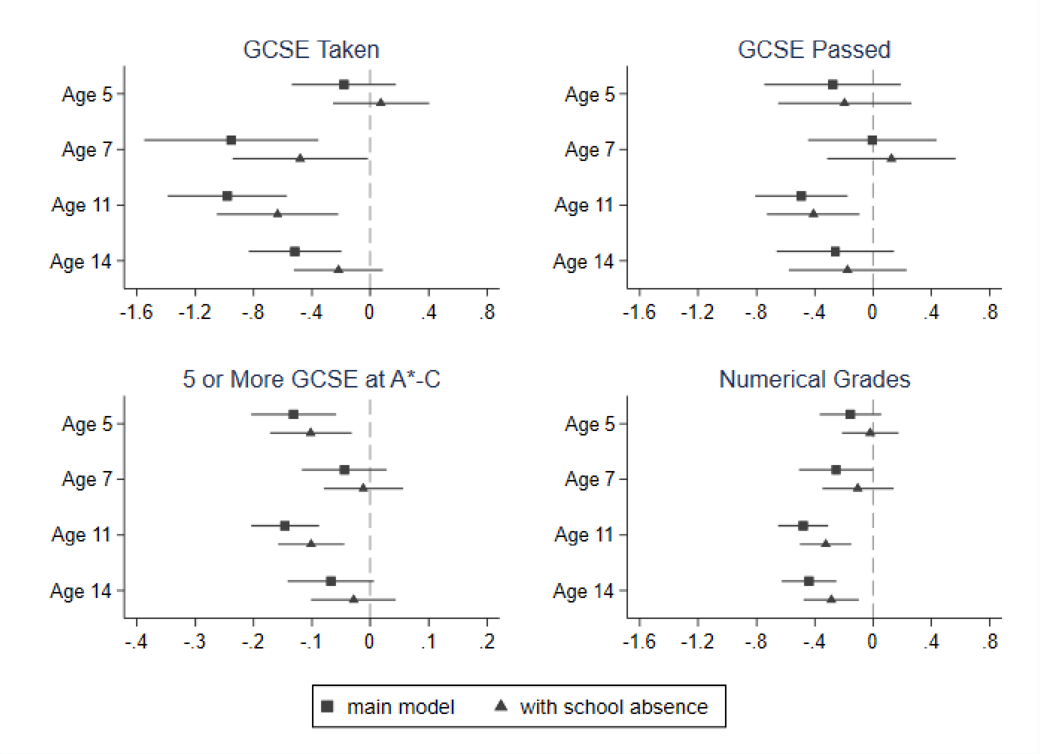

*Notes*: This figure plots the estimates from the main analysis (Table 3) and the one with the additional control of school absence (Appendix Table A4) based on the sample for whom the school attendance data are available (n=4,636).

**Appendix Table A1. Association between Emotional Problem Shocks (Measured by the SDQ Emotional Symptoms Sub-scale≥5) and GCSE Performance without Controlling for the Comorbidity of Behavioural Problems**

| | (1) Number of GCSE Subjects Taken | (2) Number of GCSE Subjects Passed | (3) Five or More Subjects with A*-C, including English and Math | (4) Average Numerical Score |
|---|---|---|---|---|
| *Emotional problems* | | | | |
| Shock at Age 5 | -0.353* | -0.462** | -0.167*** | -0.293*** |
| | (0.189) | (0.218) | (0.035) | (0.100) |
| Shock at Age 7 | -1.262*** | -0.128 | -0.078** | -0.397*** |
| | (0.344) | (0.218) | (0.037) | (0.124) |
| Shock at Age 11 | -1.138*** | -0.645*** | -0.169*** | -0.593*** |
| | (0.214) | (0.161) | (0.030) | (0.089) |
| Shock at Age 14 | -0.669*** | -0.382* | -0.088** | -0.523*** |
| | (0.170) | (0.200) | (0.039) | (0.101) |
| Observations | 4,783 | 4,783 | 4,491 | 4,718 |

*Notes*: This table reports the associations between mental health shock measured by the SDQ Emotional Symptoms Sub-scale (≥5) and GCSE outcomes. These analyses do not control for a set of health shock indicators of behavioural problems as common comorbidity. Models presented in Columns (1)-(4) all control for children's sex, ethnicity, birthweight, year and month of birth, mother's age of birth, education, maternal depression, number of siblings, number of household members and income at the baseline. Column (2) additionally controls for the number of GCSE subjects taken. Column (3) restricts the sample to children taking more than five GCSE subjects. It estimates a Probit model and presents the average marginal effects of the corresponding estimates. Column (4) restricts the sample to children with available numerical GCSE scores. Statistical significance levels are shown as *** p<.01, ** p<.05, * p<.1.

**Appendix Table A2. Association between Mental Health Shocks and School Absenteeism**

| | (1)<br>School Absence Ratio<br>Total Difficulty Score≥17 | (2)<br>School Absence Ratio<br>SDQ Emotional Sub-scale≥5 |
|---|---|---|
| Shock at Age 5 | 1.831*** | 1.349*** |
| | (0.436) | (0.380) |
| Shock at Age 7 | 2.459*** | 2.510*** |
| | (0.640) | (0.480) |
| Shock at Age 11 | 2.060*** | 1.829*** |
| | (0.388) | (0.378) |
| Shock at Age 14 | 2.054*** | 1.575*** |
| | (0.405) | (0.312) |
| Observations | 4,636 | 4,636 |

*Notes*: This table reports the associations between mental health shocks and school absence ratio. Mental health shocks are measured by the Total Difficulty Score (≥17) in Column (1) and the SDQ Emotional Sub-scale(≥5) in Column (2). Models presented in Columns (1)-(2) all control for children's sex, ethnicity, birthweight, year and month of birth, mother's age of birth, education, maternal depression, number of siblings, number of household members and income at the baseline. Column (2) additionally controls for a set of health shock indicators of behavioural problems measured by the SDQ conduct problems sub-scale (≥4) as common comorbidity. Statistical significance levels are shown as *** p<.01, ** p<.05, * p<.1.

**Appendix Table A3. Association between Mental Health Shocks (Measured by the Total Difficulty Score≥17) and GCSE Performance Controlling for School Absenteeism**

| | (1) Number of GCSE Subjects Taken | (2) Number of GCSE Subjects Passed | (3) Five or More Subjects with A*-C, including English and Math | (4) Average Numerical Score |
|---|---|---|---|---|
| *General mental health problems* | | | | |
| Shock at Age 5 | -0.894*** | -0.861*** | -0.180*** | -0.581*** |
| | (0.236) | (0.231) | (0.037) | (0.100) |
| Shock at Age 7 | -1.477*** | -0.679*** | -0.162*** | -0.747*** |
| | (0.300) | (0.250) | (0.041) | (0.117) |
| Shock at Age 11 | -1.027*** | -1.221*** | -0.207*** | -0.904*** |
| | (0.211) | (0.201) | (0.037) | (0.101) |
| Shock at Age 14 | -0.729*** | -0.871*** | -0.180*** | -0.681*** |
| | (0.230) | (0.242) | (0.041) | (0.141) |
| School Absence Rate | -0.187*** | -0.066*** | -0.025*** | -0.092*** |
| | (0.016) | (0.015) | (0.002) | (0.008) |
| Observations | 4,636 | 4,636 | 4,422 | 4,575 |

*Notes*: This table reports the associations between mental health shocks measured by the Total Difficulty Score (TDS≥17) and GCSE outcomes. Models presented in Columns (1)-(4) all control for children's sex, ethnicity, birthweight, year and month of birth, mother's age of birth, education, maternal depression, number of siblings, number of household members and income at the baseline. Column (2) additionally controls for the number of GCSE subjects taken. Column (3) restricts the sample to children taking more than five GCSE subjects. It estimates a Probit model and presents the average marginal effects of the corresponding estimates. Column (4) restricts the sample to children with available numerical GCSE scores. Statistical significance levels are shown as *** p<.01, ** p<.05, * p<.1.

**Appendix Table A4. Association between Emotional Problem Shocks (Measured by SDQ Emotional Symptoms Sub-scale≥5) and GCSE Performance Controlling for School Absenteeism**

| | (1)<br>Number of GCSE Subjects Taken | (2)<br>Number of GCSE Subjects Passed | (3)<br>Five or More Subjects with A*-C, including English and Math | (4)<br>Average Numerical Score |
|---|---|---|---|---|
| *Anxiety disorders* | | | | |
| Shock at Age 5 | -0.067 | -0.368* | -0.133*** | -0.149 |
| | (0.171) | (0.217) | (0.034) | (0.091) |
| Shock at Age 7 | -0.671*** | -0.001 | -0.041 | -0.222* |
| | (0.247) | (0.224) | (0.035) | (0.121) |
| Shock at Age 11 | -0.780*** | -0.552*** | -0.124*** | -0.447*** |
| | (0.219) | (0.165) | (0.029) | (0.093) |
| Shock at Age 14 | -0.310* | -0.284 | -0.047 | -0.373*** |
| | (0.158) | (0.207) | (0.038) | (0.102) |
| School Absence Rate | -0.195*** | -0.071*** | -0.026*** | -0.098*** |
| | (0.017) | (0.015) | (0.002) | (0.008) |
| Observations | 4,636 | 4,636 | 4,422 | 4,575 |

*Notes*: This table reports the associations between mental health shocks measured by the SDQ Emotional Symptoms Sub-scale (≥5) and GCSE outcomes. A set of health shock indicators of behavioural problems measured by the SDQ conduct problems sub-scale (≥4) is controlled for as common comorbidity. Models presented in Columns (1)-(4) all control for children's sex, ethnicity, birthweight, year and month of birth, mother's age of birth, education, maternal depression, number of siblings, number of household members and income at the baseline. Column (2) additionally controls for the number of GCSE subjects taken. Column (3) restricts the children to those taking more than five GCSE subjects. It estimates a Probit model and presents the average marginal effects of the corresponding estimates. Column (4) restricts the sample to children with available numerical GCSE scores. Heteroskedasticity-robust standard errors are used. Statistical significance levels are shown as *** p<.01, ** p<.05, * p<.1.

**Appendix Table A5. Association between Mental Health Shocks and Likelihood for Studying at the University**

| | (1) | (2) | (3) | (4) |
|---|---|---|---|---|
| | Children's Evaluation (4 categories recoded from 0 - 100) | | Parents' Evaluation (1=unlikely - 4=very likely) | |
| | Total Difficulty Score | SDQ Emotional Symptoms Sub-scale | Total Difficulty Score | SDQ Emotional Symptoms Sub-scale |
| *Mental health problems/Emotional problems* | | | | |
| Shock at Age 5 | 0.700* | 0.993 | 0.670* | 1.120 |
| | (0.138) | (0.210) | (0.144) | (0.184) |
| Shock at Age 7 | 0.413*** | 0.618** | 0.457*** | 0.938 |
| | (0.097) | (0.138) | (0.110) | (0.198) |
| Shock at Age 11 | 0.443*** | 0.593*** | 0.361*** | 0.668** |
| | (0.132) | (0.099) | (0.076) | (0.109) |
| Shock at Age 14 | 0.638* | 0.767 | 0.541** | 0.768 |
| | (0.164) | (0.138) | (0.143) | (0.125) |
| Observations | 2,835 | 2,835 | 3,397 | 3,397 |

*Notes*: This table reports the associations between mental health shocks and the perceived probability of studying at the university. Mental health shocks were measured by the Total Difficulty Score (≥17) in Columns (1) and (3) and the SDQ Emotional Symptoms Sub-scale (≥5) in Columns (2) and (4). When using the SDQ Emotional Symptoms Sub-scale (≥5), a set of health shock indicators of behavioural problems measured by the SDQ conduct problems sub-scale (≥4) is controlled for as common comorbidity. Models presented in Columns (1)-(4) all control for children's sex, ethnicity, birthweight, year and month of birth, mother's age of birth, education, maternal depression, number of siblings, number of household members and income at the baseline. Children's evaluation was originally reported using a number between 0 and 100. We converted the response into a four-category variable using 25, 50 and 75 as cut-offs. Parents' evaluation is reported as a categorical variable between 1 and 4, which is estimated using an order logit model. Columns (3) and (4) present the odds ratio. Statistical significance levels are shown as *** p<.01, ** p<.05, * p<.1.

**Appendix Table A6. Model Parameters to Calculate Productivity Loss**

| Equation | Parameter | Description | Value | Source |
|---|---|---|---|---|
| A1 | $\alpha$ | Discounted rate | 3.5% | HM Treasury[4] |
| A1 | $\beta$ | Labour income annual increasing rate | 2% | Hypothetical inflation rate |
| A2 | $p_{job}$ | Proportion of the working-age population active in the labour force | 75.02% | Official Statistics: Employment rate in 2023[5] |
| A2 | $p_f$ | Proportion of workers with full-time employment | 74.41% | Official Statistics: Full-time, part-time and temporary workers in 2023[6] |
| A2 | $h_f, h_p$ | Average weekly working hours of a full-time and part-time worker | $h_f = 36.67$, $h_p = 16.47$ | Official Statistics: Actual weekly hours worked in 2023[7] |
| A2 | $rate$ | Hourly rate (for individuals without the GCSE qualification) | £10.42 | UK minimum wage in 2023 for 23 and over[8] |

### References

1. Department for Education. GOV.UK [Internet]. 2020 [cited 2024 Sep 13]. Level 2 and 3 attainment by young people aged 19 in 2019. Available from: https://www.gov.uk/government/statistics/level-2-and-3-attainment-by-young-people-aged-19-in-2019

2. Department for Education. GOV.UK [Internet]. 2016 [cited 2024 Sep 9]. GCSE and A level differences in England, Wales and Northern Ireland. Available from: https://www.gov.uk/government/publications/gcse-and-a-level-differences-in-england-wales-and-northern-ireland

3. Mcintosh S. Further Analysis of the Returns to Academic and Vocational Qualifications. Oxford Bulletin of Economics and Statistics. 2006;68(2):225–51. doi:10.1111/j.1468-0084.2006.00160.x

4. HM Treasury. GOV.UK [Internet]. 2022 [cited 2024 Dec 2]. Green Book supplementary guidance: discounting. Available from: https://www.gov.uk/government/publications/green-book-supplementary-guidance-discounting

5. ONS. Employment in the UK: September 2024 [Internet]. 2024 [cited 2024 Dec 2]. Employment in the UK - Office for National Statistics. Available from: https://www.ons.gov.uk/employmentandlabourmarket/peopleinwork/employmentandemployeetypes/bulletins/employmentintheuk/september2024

6. ONS. Full-time, part-time and temporary workers (seasonally adjusted) - Office for National Statistics [Internet]. 2024 [cited 2024 Dec 2]. Available from: https://www.ons.gov.uk/employmentandlabourmarket/peopleinwork/employmentandemployeetypes/datasets/fulltimeparttimeandtemporaryworkersseasonallyadjustedemp01sa

7. ONS. Actual weekly hours worked (seasonally adjusted) - Office for National Statistics [Internet]. 2024 [cited 2024 Dec 2]. Available from: https://www.ons.gov.uk/employmentandlabourmarket/peopleinwork/earningsandworkinghours/datasets/actualweeklyhoursworkedseasonallyadjustedhour01sa

8. GOV.UK. National Minimum Wage and National Living Wage rates [Internet]. 2024 [cited 2024 Dec 2]. Available from: https://www.gov.uk/national-minimum-wage-rates
